## Supplementary material for "A postzygotic *GNA13* variant upregulates the RHOA/ROCK pathway and alters melanocyte function in a mosaic skin hypopigmentation syndrome": Table 1

| Patient | | Patient 1 | Patient 2 | Patient 3 | Patient 4 |
|---|---|---|---|---|---|
| ***GNA13* Variant** | **Nomenclature** | chr17:g.63010910C>T<br>NM_006572.4:c.599G>A<br>p.(Arg200Lys) | chr17:g.63010910C>T<br>NM_006572.4:c.599G>A<br>p.(Arg200Lys) | chr17:g.63010910C>T<br>NM_006572.4:c.599G>A<br>p.(Arg200Lys) | chr17:g.63010910C>T<br>NM_006572.4:c.599G>A<br>p.(Arg200Lys) |
| **Tissue VAF*** | **Affected skin** | 29.0% | 30.0% | 25.0% | 36% |
| | **Blood** | 0.3% (NS) | 0.00% (NS) | - | 0.0% (NS) |
| | **Buccal swab** | 0.1% (NS) | 10.6% | 3.9% | 0.5% (NS) |
| | **Urine** | 0.0% (NS) | - | 0.1% (NS) | 0.0% (NS) |
| **Sex** | | Male | Male | Female | Female |
| **Age at last visit** | | 11-15 | 21-25 | 0-5 | 6-10 |
| **Growth** | **OFC (cm)** | 50.5 (-1 SD) | - | - | - |
| | **Height (cm)** | 108.5 (-0.5 SD) | - | - | 123 (+ 0.5 SD) |
| | **Weight (kg)** | 19.0 (M) | - | - | 22 (M) |
| **Asymmetry**<br>**Craniofacial anomalies** | | Right-sided hemihypotrophy (lower limb) | Right-sided hemifacial hypoplasia<br>Lower limb asymmetry<br>Plagiocephaly<br>Scoliosis | No | Right-sided hemifacial hypoplasia |
| **Pregnancy and birth** | **Fetal ultrasonography** | - | Intrauterine growth retardation | - | Normal |
| | **Term** | 41 wk | - | - | 39 wk |
| | **OFC at birth (cm)** | NR | - | - | 33.5 |
| | **Length at birth (cm)** | 44.0 | - | - | 49.5 |
| | **Weight at birth (g)** | 2500 | - | - | 3560 |
| **Skin and hair** | **Hypopigmentation pattern** | Linear, spindle-shaped, flag-like, splash-like, phylloid | Linear, spindle-shaped | Linear, splash-like | Linear, spindle-shaped |
| | **Location of hypopigmentation** | right upper and lower limbs, trunk | Lower limbs bilaterally, face (chin) | Left lower limb | Right upper limb |
| | **Hair anomalies** | Absence of hair on hypopigmented skin | Patchy alopecia of scalp hair and right eyelashes | NR | Patchy alopecia on parietal scalp and right eyelashes |
| | **Wound healing** | Delayed wound closure (9 months) after skin biopsy (right upper limb) | Delayed wound closure after skin biopsy (1 month) and after excision of supernumerary digit of the right hand (2 - 3 months) | NR | NR |
| **Neurology** | **Peripheral nerves** | Right foot dysaesthesia and weakness | NR | NR | NR |
| | **Brain MRI** | White matter hyperintensities (centrum semiovale, right parietal lobe and temporoparietal junction, left frontal lobe) | Hydrocephalus | Normal (at age 6 mo.) | Not performed |
| **Acral anomalies** | | Camptodactyly | Right hand polysyndactyly<br>Left hand brachymetacarpy<br>Bilateral pes planus (partial talocalcaneal fusion)<br>Toe syndactyly (right foot) | Left foot polysyndactyly | No |
| **Ocular anomalies** | | No | Coloboma of upper eyelid, bilateral abnormal fundus | Mild iris coloboma | Amblyopia with mild microphalmia of right eye<br>Narrow palpebral fissure of right eye<br>Normal eye fundus |
| **Dental anomaly** | | No | Odontogenic cysts, dental anomalies | NR | Conical teeth |
| **Hearing loss** | | No | Mixed hypoacusis of right ear (hearing aid)<br>Otosclerosis<br>Abnormal inner auditory and cochlear nerve canals<br>Cochlear nerve agenesis or hypoplasia | Left ear hypoacusis<br>Chronic otitis media | No |
| **Gastrointestinal tract anomalies** | | No | Jejunal atresia | Colonic atresia (ascending colon) | Colonic atresia |
| **Urinary tract anomalies** | | No | Bilateral hydronephrosis, right megaureter | No | No |

VAF: variant allele frequency
* Targeted ultradeep sequencing (TUDS) on affected skin
NR : not reported
(-) missing data
NS: not significant (below background noise threshold)
